# Individual Reference Intervals for Personalized Interpretation of Clinical and Metabolomics Measurements

**DOI:** 10.1101/2021.06.04.21258334

**Authors:** Murih Pusparum, Gökhan Ertaylan, Olivier Thas

## Abstract

The Population Reference Interval (PRI) refers to the range of outcomes that are expected in a healthy population for a clinical or a diagnostic measurement. This interval is widely used in daily clinical practice and is essential for assisting clinical decision making in diagnosis and treatment. In this study, we demonstrate that each individual indeed has a range for a given variable depending on personal biological traits. This Individual Reference Intervals (IRI) can be calculated and be utilized in clinical practice, in combination with the PRI for improved decision making where multiple data points are present per variable. As calculating IRI requires several data points from the same individual to determine a personal range, here we introduce novel methodologies to obtain the correct estimates of IRI. We use Linear Quantile Mixed Models (LQMM) and Penalized Joint Quantile Models (PJQM) to estimate the IRI’s upper and lower bounds. The estimates are obtained by considering both the within and between subjects’ variations. We perform a simulation study designed to benchmark both methods’ performance under different assumptions, resulted in PJQM giving a better empirical coverage than LQMM. Finally, both methods were evaluated on real-life data consisting of eleven clinical and metabolomics parameters from the VITO IAM Frontier study. The PJQM method also outperforms LQMM on its predictive accuracy in the real-life data setting. In conclusion, we introduce the concept of IRI and demonstrate two methodologies for calculating it to complement PRIs in clinical decision making.

## Introduction

In the early days of clinical practice diagnostic, (laboratory) tests were developed to be performed at the presence of the patient with basic equipment, resulting in rapid evaluation of the results, and the rendering of a diagnostic opinion. Choice of an appropriate test, its performance, and interpretation entirely depended on the practitioner.^1^

In today’s medical practice, clinical laboratory tests are routinely performed by certified clinical laboratories for examining the clinical, physiological or molecular state of a patient. Reference intervals (RI) are essential for the interpretation of clinical laboratory tests, assisting the professionals regarding diagnosis and decision making in patient care. Such intervals are calculated from a reference population of healthy individuals and will be referred as *Population Reference Intervals* (PRI).^2^ For any particular clinical parameter, the specific PRI is compared with the test measurement from the patient; when the measurement is within the PRI a normal reading is declared and no further action is needed. If the test measurements are outside the PRI boundaries, the context of the patient explaining the result and other test measurements are taken into account to determine a course of action.

At present, the PRI estimation methods^3,4,5^ typically require cross-sectional data, i.e. data that consist of one measurement for each subject in the sample. The latter is assumed to represent a population of healthy people. In general, the methods can be grouped into the parametric and the non-parametric methods. The former relies on distributional assumptions, while the latter simply estimates the PRI bounds by the appropriate sample order statistics (i.e. the empirical quantiles). Throughout this paper, these methods will be referred to as the classical PRI estimation methods. In practice, many PRIs are computed using the non-parametric methods as they are free of distributional assumptions. Combined with the bootstrap resampling technique, it may provide a robust estimate and it fulfills the recommendations of the International Federation of Clinical Chemistry.^6,7^ A study has been presented to compare these classical methods for PRI estimations on a large cross-sectional data.^2^ These methods, however, are not appropriate when data comes as time-series obtained from the same subject as they do not adequately capture the within and between-subjects variability.

Despite the widespread use of the PRI in daily clinical practice, it falls short in providing the individual context necessary to recognize diseases at an early stage. Precision health starts from the premise that each person has its individual specification of “healthy", resulting from individual biological traits. From this perspective, definition of personalised health would benefit from PRIs that are subject-specific. We refer to such intervals as *Individual Reference Intervals* (IRI). Such an interval would indicate what test results can be expected with e.g. a probability of 95%, when that person is in its healthy state. In this paper, we will propose a method for the estimation of IRIs, starting from time-series data with multiple measurements for each subject.

In this paper, two estimation procedures are described for constructing the IRIs, allowing for both variations between and within subjects, and requiring time-series data for model fitting. The first approach makes use of Linear Quantile Mixed Models (LQMM) and has been recently proposed,^2^ but it has not been properly evaluated yet. Since this method only allows for the separate estimation of the lower and upper bounds of the IRIs, we will refer to them as the *separate IRIs*. Our second method is based on a new Joint Quantile Model (JQM) that simultaneously models the lower and upper bounds. A penalized parameter estimation is proposed, which allows for the calculation of IRIs even when only 5 repeated outcomes per subject are available. These IRIs will be referred to as the *joint IRIs*.

In Section *Models and Methods*, we describe the methods and the model formulations. In Section *Simulation Study*, the methods are evaluated, both using simulated data as well as the real-life cohort study data. This cohort study data has been collected by The Flemish Institute for Technological Research (VITO) in a pilot time-series study in which, during 12 months, 30 healthy individuals donated blood, urine, and stool samples at monthly visits.^8^ At bimonthly visits, both their clinical and metabolomics outcomes were measured and assessed by accredited labs and appointed doctors. Finally, a conclusion is formulated in Section *Discussion and Conclusion*.

## Models and Methods

### Linear Quantile Mixed Model

The term *separate-IRI* is used for an IRI for which the two IRI boundaries are separately estimated. In this section we describe a method based on fitting two LQMMs.^9,10^ We formulate two LQMMs that only include a fixed intercept and a subject-specific random intercept to model the between-subject variability of the reference intervals.

Consider the quantile function *Q*(*τ*) of a random variable *Y*. This is defined for all *τ* ∈ [0, 1] as *Q*(*τ*) = inf {*y* ∈ ℝ : *F* (*y*) ≥ *τ*}. The index *i* is added to the quantile function, i.e. *Q*_*i*_(*τ*), to make it refer to the distribution of the outcome *Y*_*i*_ of subject *i* = 1, …, *n*. The probabilities *τ*_1_ and *τ*_2_ *> τ*_1_ are chosen so as to make [*Q*_*i*_(*τ*_1_), *Q*_*i*_(*τ*_2_)] the IRI of subject *i* with nominal coverage probability *τ*_2_ − *τ*_1_. The Quantile model for subject *i* then becomes

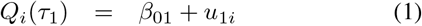

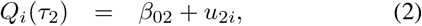

where *β*_01_, *β*_02_ ∈ ℝ are the fixed intercepts, and *u*_1*i*_ and *u*_2*i*_ are the subject-specific random effects. In this paper we assume these random intercepts *u*_1*i*_ and *u*_2*i*_ to follow zero-mean normal distributions with variances *ψ*_*u*1_ and *ψ*_*u*2_, respectively. Moreover, the random intercepts are assumed to be independently distributed.

For the estimation of the parameters, we follow the procedure of quantile regression in longitudinal data with only a random intercept.^9,10^ Let *y*_*ij*_ denote the *j*th outcome (e.g. clinical parameter of interest) on subject *i* (*i* = 1, …, *N, j* = 1, …, *n*_*i*_). As a working model, the estimation procedure makes the assumption that the outcomes *y*_*ij*_ on the *i*th subject, conditionally on *u*_*ki*_, are independently distributed according to the Asymmetric Laplace Distribution (ALD) with density function (*k* = 1, 2)^9^

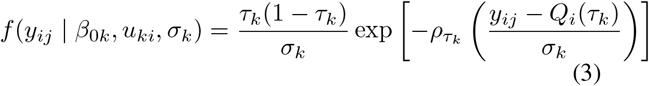

where *Q*_*i*_(*τ*_*k*_) = *β*_0*k*_ + *u*_*ki*_ is the model for the *τ*_*k*_th quantile. The parameter *σ*_*k*_ is the ALD scale parameter and 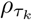 is the check function for the *τ*_*k*_-th quantile. A range of estimation strategies have been established for fitting this model;^9,10^ we will use the implementation in the *lqmm* R package.^11^

### Joint Quantile Model

#### Model Description

In this section we introduce a Joint Quantile Model (JQM) which forms the basis for simultaneously estimating the bounds of the IRIs. To distinguish these intervals from the separate IRIs, we will use the term *joint-IRI* to refer to the IRIs arising from the JQM. Upon using the same notation as before, we define the JQM by the following three equations (*i* = 1, …, *N* ; *j* = 1, …, *n*_*i*_):

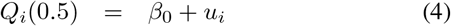

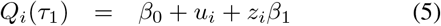

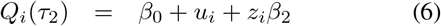

where *β*_0_ and the subject-specific random effects *u*_*i*_ are as before, except that they are now shared with all models (hence the term *joint* quantile model). According to this model the median of the outcome of subject *i* is given by *β*_0_ + *u*_*i*_. The model will need a restriction on these parameters to make them identifiable (see later). Similar as with the separate-IRI, when we assign *τ*_1_ and *τ*_2_ *> τ*_1_ such that *τ*_1_ + *τ*_2_ = 1, we can interpret the quantile function in (5) and (6) as the lower and the upper bounds of an IRI for subject *i*, with coverage probability *τ*_2_ — *τ*_1_. The length of the IRI is given by *Q*_*i*_(*τ*_2_) − *Q*_*i*_(*τ*_1_) = *z*_*i*_(*β*_2_ − *β*_1_). Hence, this model does not only allow for subject-specific locations of the reference intervals (given by *u*_*i*_), but it also allows for subject-specific lengths. In this respect, the *z*_*i*_s are individual scaling factors restricted to be non-negative (*z*_*i*_ ≥ 0), and *β*_0_ + *β*_1_ and *β*_0_ + *β*_2_ are the lower and upper bounds of an IRI of a subject with *u*_*i*_ = 0 and *z*_*i*_ = 1.

Since the *β*-parameters are shared in three quantiles, these parameters will need to be estimated by considering the three models simultaneously. Several approaches for estimating the model parameters can be considered. For example, we could assume that the subject-specific *u*_*i*_ and *z*_*i*_ are random effects, distributed according to a user-specified distribution (e.g. normal distribution for *u*_*i*_ and inverse gamma for *z*_*i*_). From this perspective, the *u*_*i*_ act as random intercepts such as in the LQMM. However, the terms *z*_*i*_*β*_1_ and *z*_*i*_*β*_2_ with *z*_*i*_ as a random effect, do not fit into the class of LQMMs. Moreover, in the LQMM approach the distribution of the random effects would need to be specified and therefore the method would enforce a strong distributional assumption which we want to avoid. While the LQMMs often focus on the inference of the model parameters, our objective is to construct IRIs that have the correct probabilistic interpretation when applied to new test outcomes of subjects. For these reasons, we do not follow the LQMM approach. Instead we propose a penalized parameter estimation procedure, which shares some characteristics with LQMM. These similarities will be discussed later. We refer the JQM method estimated by this penalized procedure as the Penalized JQM (PJQM).

#### Parameter Estimation

We propose to estimate the *β*_*k*_, *k* = 1, 2, and the subject-specific effects by minimizing the function

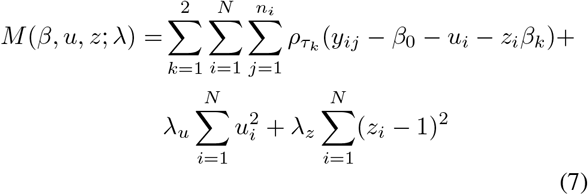

in which *ρ*_*τ*_ (*w*) = *w*(*τ* — *w* ≤ 0) is the check function. The first term corresponds to the objective function for estimating parameters in linear quantile models, and the two last terms are penalty terms with user-specified penalty parameters *λ*_*u*_, *λ*_*z*_ ≥ 0. The first penalty term forces the subject-specific effects *u*_*i*_ to zero as *λ*_*u*_ increases (i.e. the subject-specific medians are shrunk towards one another), and the scaling factors *z*_*i*_ are shrunk towards 1 as *λ*_*z*_ increases, bringing the lengths of the IRIs closer to one another.

The function *M* can also be considered as a log-likelihood function of a LQMM. The penalty terms then arise if the *u*_*i*_ and *z*_*i*_ were considered as normal random effects with mean zero and one, respectively, and the *λ* penalty parameters then correspond to the (inverse) variance parameters. However, the theory of LQMMs do not accommodate for terms of the form *z*_*i*_*β*_*k*_,^9,10^ and our focus is not on statistical inference. Our motivation is twofold. First, our model specification requires restrictions on the parameters to make them identifiable. This is accomplished by the penalization of the *u*_*i*_s and *z*_*i*_s. Second, the penalization terms (or shrinkage) cause information sharing between subjects. This is necessary, because in realistic applications the numbers of replicates per subject (*n*_*i*_) are quite small, too small to nonparametrically estimate e.g. the 2.5% and 97.5% quantiles of IRIs. The user-specified penalty parameters will be selected by means of a cross-validation procedure aiming at calibrating the IRIs at the nominal coverage probability *τ*_2_ − *τ*_1_.

In order to minimize the objective function for a given *λ*_*u*_ and *λ*_*z*_, and obtain the estimates of all model parameters, we applied an iterative procedure. This iterative procedure is explained more in detail in Section 1 in the *Supplementary Document*. The procedure for selecting the penalty parameters is described in Section *Selection of the Penalty Parameters*. First we need to show how IRIs are computed for new subjects, i.e. for a subject that did not contribute its data for the estimation of the parameters.

#### IRI for a New Subject

Suppose we have estimates of all *β* parameters and of all *u*_*i*_s and *z*_*i*_*s*. The latter only refer to subjects with data present in the dataset. We now aim to compute the IRI of a new subject, say subject *m*. We can still use the estimates 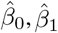 and 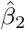, but we need estimates of the new subject-specific effects *u*_*m*_ and *z*_*m*_. Our method proceeds along the following steps:

1. Set 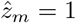 (as if the new subject is an *average*
2. Estimate *u*_*m*_ by minimising

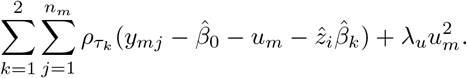 The solution is denoted by 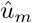
3. Estimate *z*_*m*_ by minimising

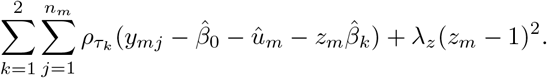 The solution is denoted by 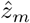
4. Iterate over the two previous steps until convergence.

The IRI of subject *m* is then given by

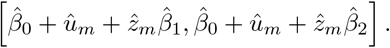

#### Selection of the Penalty Parameters

The selection of penalty parameters proceeds along the lines of a leave-one-out cross validation scheme. Consider two sets of possible values for *λ*_*u*_ and *λ*_*z*_, say *L*_*u*_ and *L*_*z*_. For each *λ*_*u*_ ∈.*L*_*u*_ and each *λ*_*z*_ ∈*L*_*z*_, the following steps are performed for each subject *i* = 1, …, *N*.

1. For a given subject *i*, split the dataset into three datasets:
  - Dataset 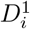: dataset with all data from subject *i*
  - Dataset 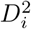: dataset with all data from the last clinical test result (except for subject *i*), i.e. the observations *y*_*ln*_*l, l* ≠ *i*.
  - All remaining data. This dataset is referred to as the *training dataset*.
2. Use the training data for estimating all parameters.
3. Compute the empirical coverage based on the observations in 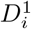:. This is calculated as the relative frequency of observations in 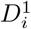: that are contained in the IRI for subject *i* (since subject *i* was not part of the training data, its IRI is calculated at outlined in Section *IRI for a New Subject*).
4. Compute the empirical coverage based on the observations in 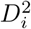.. This is calculated as the relative frequency of observations in 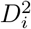 that are contained in the IRIs of the corresponding subjects.
5. Calculate the overall empirical coverage as the average of the two previous coverages.

Upon completion of all these steps for all subjects *i* = 1, …, *N*, the total empirical coverage is calculated as the average overall empirical coverage over all subjects. The optimal (*λ*_*u*_, *λ*_*z*_) is then the parameter combination that resulted in the smallest total empirical coverage that is at least as large as the nominal coverage level.

## Simulation Study

### Simulated Data

We conducted a simulation study to evaluate the performance of the LQMM and the Penalized JQM (PJQM) methods. Data were generated with linear mixed models (LMM),

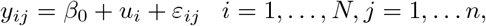

with *β*_0_ fixed at 0 and the *u*_*i*_ are random intercepts. We considered both standard normal distribution and a scaled 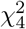 distribution for simulating the *u*_*i*_. The latter is scaled so that is has mean 0 and variance 1. Since both the LMM and the quantile models (LQMM and PJQM) are location-shift models, *β*_0_ and the *u*_*i*_s from the LMM correspond to the *β*_0_ and the *u*_*i*_s of our quantile models. For the error terms we considered a normal distribution with mean zero and variance 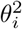. This variance was either set to a constant 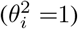. such as in the LQMM method scenario, or it was sampled from an inverse gamma distribution with shape parameter *a* and scale parameter *b*. The latter generates outcomes with subject-specific variances (heteroscedasticity). A scaled *t*_3_ distribution was also considered for representing a symmetric distribution with heavier tails than the normal distribution. Table S1 in the *Supplementary Document* gives a description of all simulation scenarios. The choice of *a* and *b* values were derived from several VITO IAM Frontier clinical outcomes and hence represents realistic settings.

For each scenario, 1000 Monte Carlo simulation runs were performed. For each simulation run, data for *N* + 1 subjects with *n* + 1 repeated outcomes were simulated. The data for the first *N* subjects and their first *n* repeated outcomes were used for estimating the model parameters. The data for the *N* + 1th subject and for the (*n* + 1)th repeated outcome were used for the estimation of the empirical coverages. The generation of data for an additional subject and for an additional time point corresponds to two possible applications in real life settings: 1) when existing subjects have new measurements, and 2) when there is a new subject that was not included in the data used for the parameter estimation. We refer the empirical coverage that corresponds to the first case as the *Time Empirical Coverage* (TEC), and to the second as the *Subject Empirical Coverage* (SEC). The average of TEC and SEC was also computed and is referred to as the *Overall Empirical Coverage* (OEC).

Figure 1 shows the OEC for all scenarios and for three different nominal coverages of the intervals. Overall, the OEC of the joint-IRIs are always larger than those of the separate-IRIs. For all numbers of repeated outcomes *n* and numbers of subjects *N*, the OEC for the joint-IRIs are consistently close to their nominal coverage levels. The OEC for the separate-IRIs, on the other hand, are always smaller than the nominal coverage, especially when *n* is small. A method with a good performance would have an OEC that is equivalent to this true nominal coverage. We also observe that the different number of subjects and random effect distributions do not strongly affect the method’s performance.

**Figure 1.**
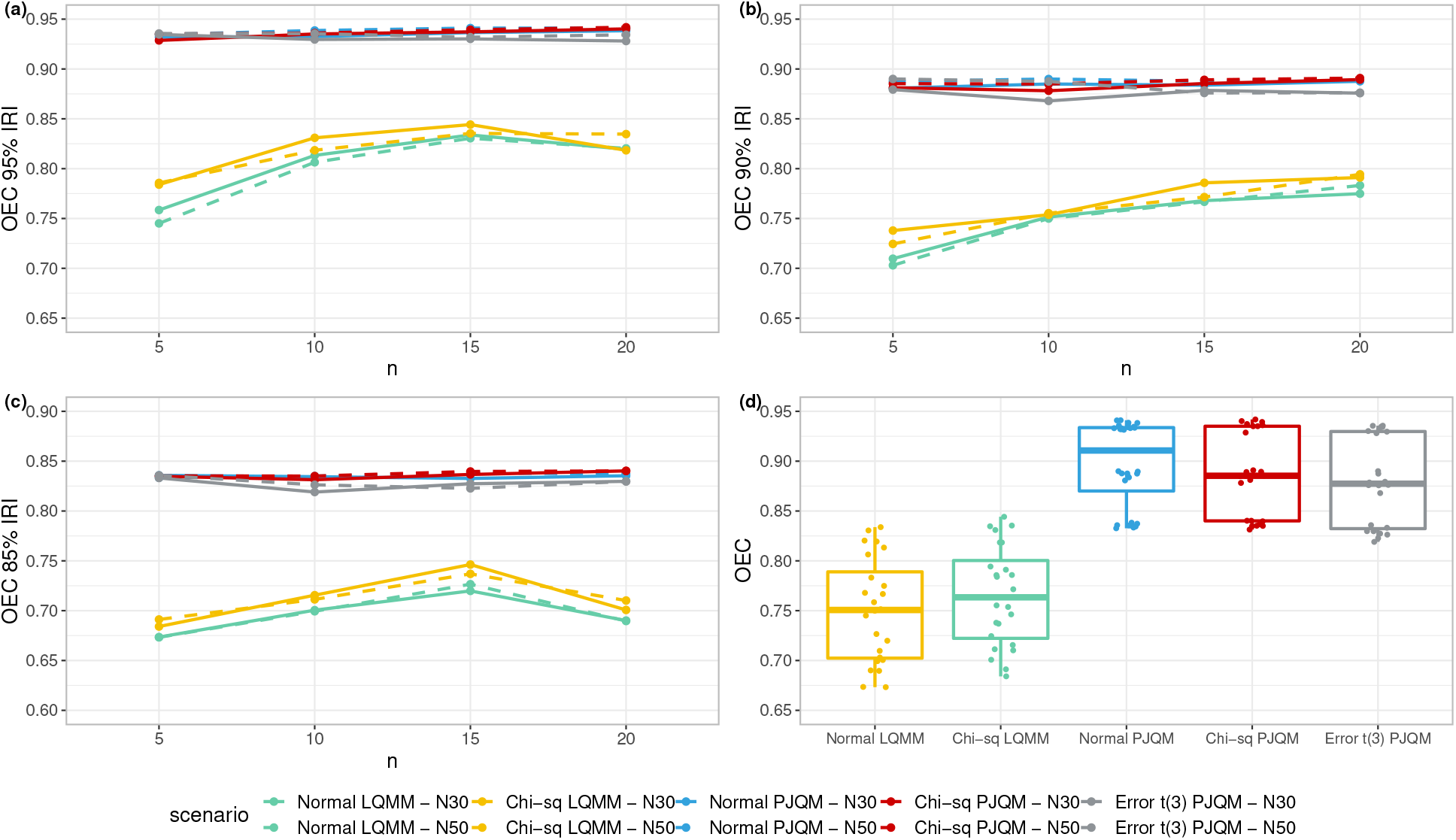
Method performance and empirical coverage distribution. The OEC were calculated for all scenarios of four different number of repeated outcomes *n* at **(a)**: 95% coverage, **(b)**: 90% coverage, and **(c)**: 85% coverage. In **(d)**: Boxplots of OEC across two methods in different random intercept and error distributions. The green, yellow, blue, and red lines correspond to scenarios with normal error terms but varying random effects’ distributions, while the grey lines correspond to scenarios with error terms follow *t*_3_ and normal random effects. In general, the OEC of joint-IRIs are always larger than for the separate-IRIs.

We have also evaluated the performance of the PJQM method by varying the parameters *a* and *b* in the data generation. The results are presented in Table 1. The number of subjects (*N*), the number of repeated outcomes within subjects (*n*), and the nominal levels were fixed to 30, 10, and 95%, respectively. The subject empirical coverages (SEC) and the overall empirical coverages (OEC) are generally close to the nominal level for all choices of *a* and *b*, suggesting the robustness of our PJQM method. However, the time empirical coverages (TEC) are somewhat smaller. The case that corresponds to the TEC calculation i.e. when existing subjects have new measurements, is presumed to affect the coverage value. In this case, the new measurements are not included in the IRI estimation procedure, resulting in a lower chance of falling within the interval. On the other hand, the SEC calculation involves both the estimates from existing subjects and the inclusion of the new subject in the IRI estimation. In Table S2 in the *Supplementary Document*, we also present the estimates penalty parameters *λ*_*u*_ and *λ*_*z*_.

**Table 1.** Performance of the PJQM method for different values of *a* and *b*. All scenarios were evaluated at 95% nominal coverage level in 30 subjects with 10 repeated outcomes.

| $a$ | $b$ | SOC | TEC | OEC |
| --- | --- | --- | --- | --- |
| 2 | 0.4 | 0.955 | 0.909 | 0.932 |
| 2 | 0.8 | 0.957 | 0.914 | 0.935 |
| 2 | 1.6 | 0.948 | 0.917 | 0.933 |
| 2.4 | 0.4 | 0.958 | 0.907 | 0.933 |
| 2.4 | 0.8 | 0.957 | 0.909 | 0.933 |
| 2.4 | 1.6 | 0.952 | 0.911 | 0.932 |
| 2.8 | 0.4 | 0.957 | 0.905 | 0.931 |
| 2.8 | 0.8 | 0.953 | 0.912 | 0.932 |
| 2.8 | 1.6 | 0.953 | 0.914 | 0.934 |

### VITO IAM Frontier Data

The VITO IAM Frontier study contains several types of outcomes, including clinical, omics (proteomics, metabolomics, genomics), and physical characteristics of 30 healthy individuals that were measured throughout a one year period. In this paper we focus on eleven outcomes from the clinical and the metabolomics measurements, assessed by two independent accredited laboratories (See Section 4 in the *Supplementary Document* for descriptions). These outcomes include glucose measurements, triglyceride, total cholesterol, LDL, HDL, and non-HDL cholesterol, albumin, creatinine, apolipoprotein A1, apolipoprotein B, and the ratio of apolipoprotein B to apolipoprotein A1. The similarity of each outcome between the clinical and metabolomics measurements is very high since in principle they measure the same chemical compound presents in the body. Although, a reasonable difference was still observed as different technologies were used. A general overview of the data and a similarity matrix are presented in Figure S1 and S2 in the *Supplementary Document*.

The performance of the LQMM and PJQM methods was investigated by computing the empirical coverages for each outcome. In particular, these empirical coverages were computed as follows: (1) all data except the data of one subject and each subject’s last observation, were used for the estimation of the model parameters; (2) the left-out data were used for the calculation of the empirical coverages (SEC, TEC and OEC). This procedure is valid because all subjects are considered healthy. In this way, the results presented in this section can be considered as a realistic, empirical assessment of the coverages. At the same time we use the data to illustrate the use of the IRIs.

We have calculated the 95% separate-IRIs and joint-IRIs for all eleven outcomes in the clinical and metabolomics data sets, measured in six time-points from bimonthly visits.

In Figure 2, we show the separate-IRIs and the joint-IRIs of glucose in the metabolomics dataset. The graph shows that the IRIs vary between subjects and that the lengths of the intervals are generally smaller for the IRIs than for the published PRI, suggesting that IRIs give more precise information for individuals than the PRI. The IRIs of triglyceride are shown in Figure 3. Note that the PRI only gives an upper bound, because a lower bound is not considered to be of medical relevance.^12^

**Figure 2.**
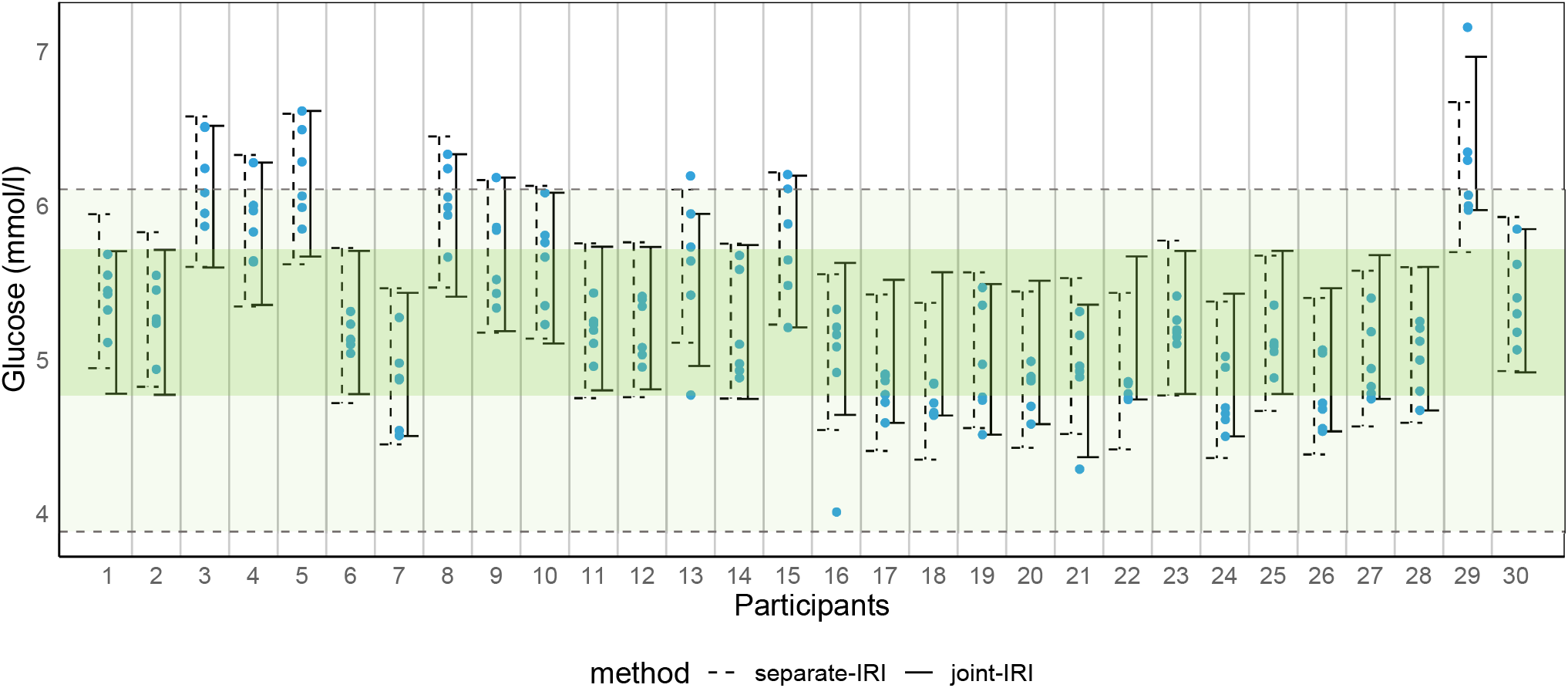
The 95% separate-IRIs and joint-IRIs for all 30 subjects for glucose measurements in the VITO IAM Frontier metabolomics data set. The blue dotted points indicate the observed measurements and the green transparent area indicates the published PRI.^12^ The darker green transparent area is the ‘average’ joint-IRI i.e. when *u*_*i*_ = 0 and *z*_*i*_ = 1 in the JQM. The order of individuals is randomised to ensure the confidentiality.

**Figure 3.**
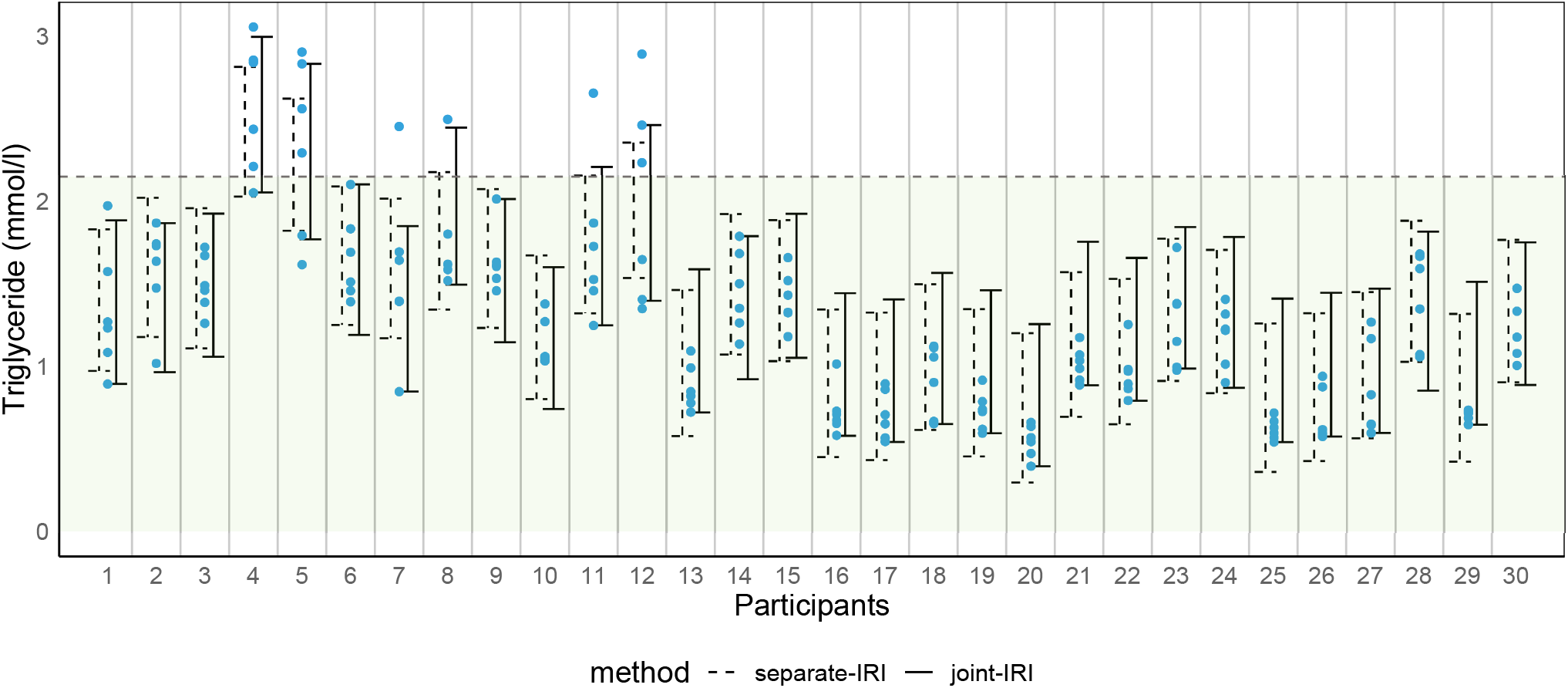
The 95% separate-IRIs and joint-IRIs for all 30 subjects for triglyceride measurements in the VITO IAM Frontier metabolomics data set. The blue dotted points indicate the observed measurements and the green transparent area indicates the published PRI.^12^ The order of individuals is randomised to ensure the confidentiality.

Figures 2 and 3 illustrate that the lengths the separate IRIs of some individuals are greater than for the joint-IRIs. The distributions of these lengths are shown as boxplots in Figure 4. These bloxplots illustrate four situations. For glucose the lengths of the joint-IRIs are generally smaller than for the separate-IRIs, while the coverage of the former is better. The joint-IRIs can thus be more personalised, while still having the correct protection against false positive results. Triglyceride is an example for which there is hardly a difference between the two methods. For total and LDL cholestoral the length distributions are about the same, but only the coverages of the joint-IRIs are close to the nominal level. Finally, for apolipoprotein B and the ratio of apolipoprotein B/A1, the lengths of the joint-IRIs are larger than those of the separate-IRIs, but the coverages of the latter are too small. Figure S3 in the *Supplementary Document* presents the IRI length distributions for other outcomes.

**Figure 4.**
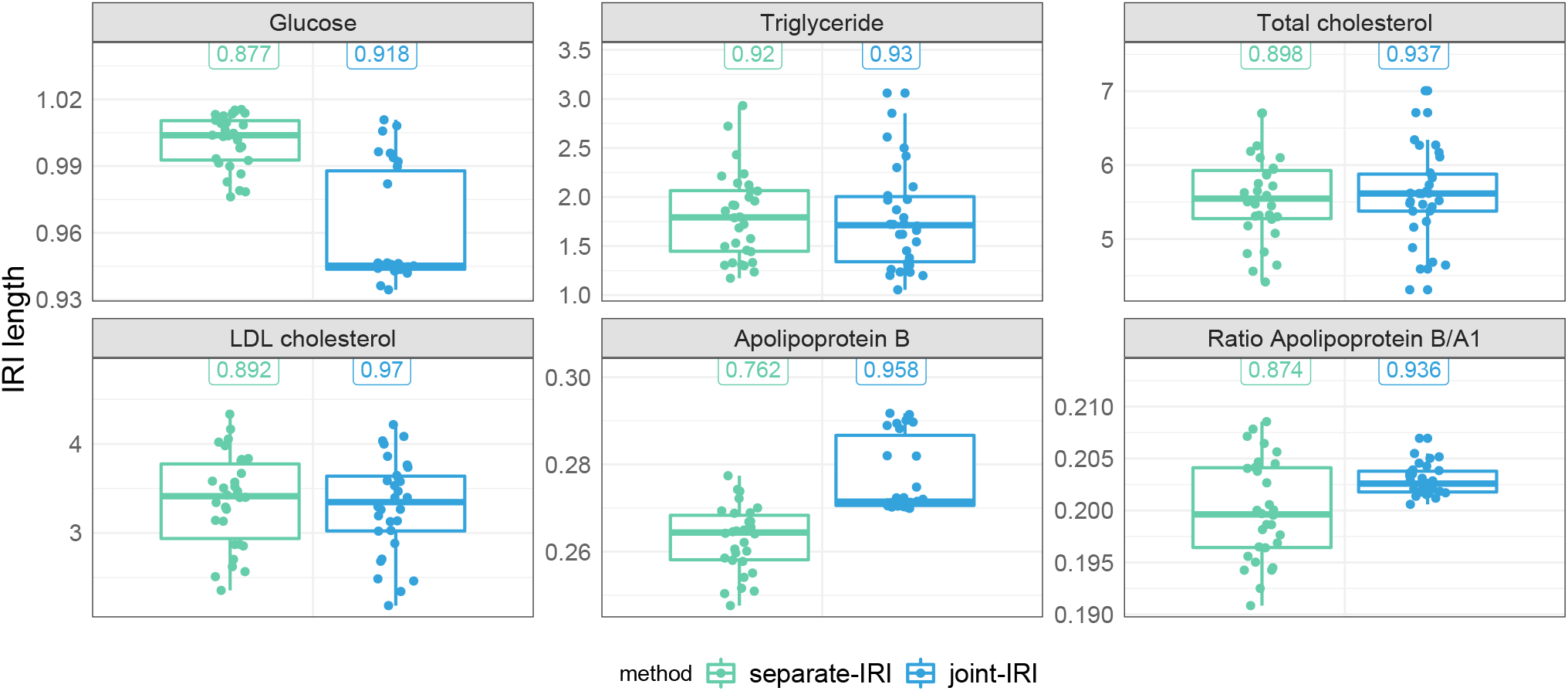
The IRI length distributions of 6 outcomes measured in the VITO IAM Frontier metabolomics data. The green and blue boxplots refer to IRI length estimated by the LQMM method (separate-IRI) and the PJQM method (joint-IRI), respectively. The values on top of each boxplot refer to the corresponding OECs. The joint IRI generally gives lower IRI length with smaller variablity.

Table 2 presents the parameter estimates and the empirical coverages for four clinical and metabolomics measurements, computed by both the LQMM and the PJQM methods at the 95% nominal level. An overall empirical coverage of as small as 69% is observed for the separate-IRIs, whereas the empirical coverages of the joint-IRIs are very close to the nominal level of 95%. Thus the joint-IRIs clearly outperform the separate-IRIs.

**Table 2.**
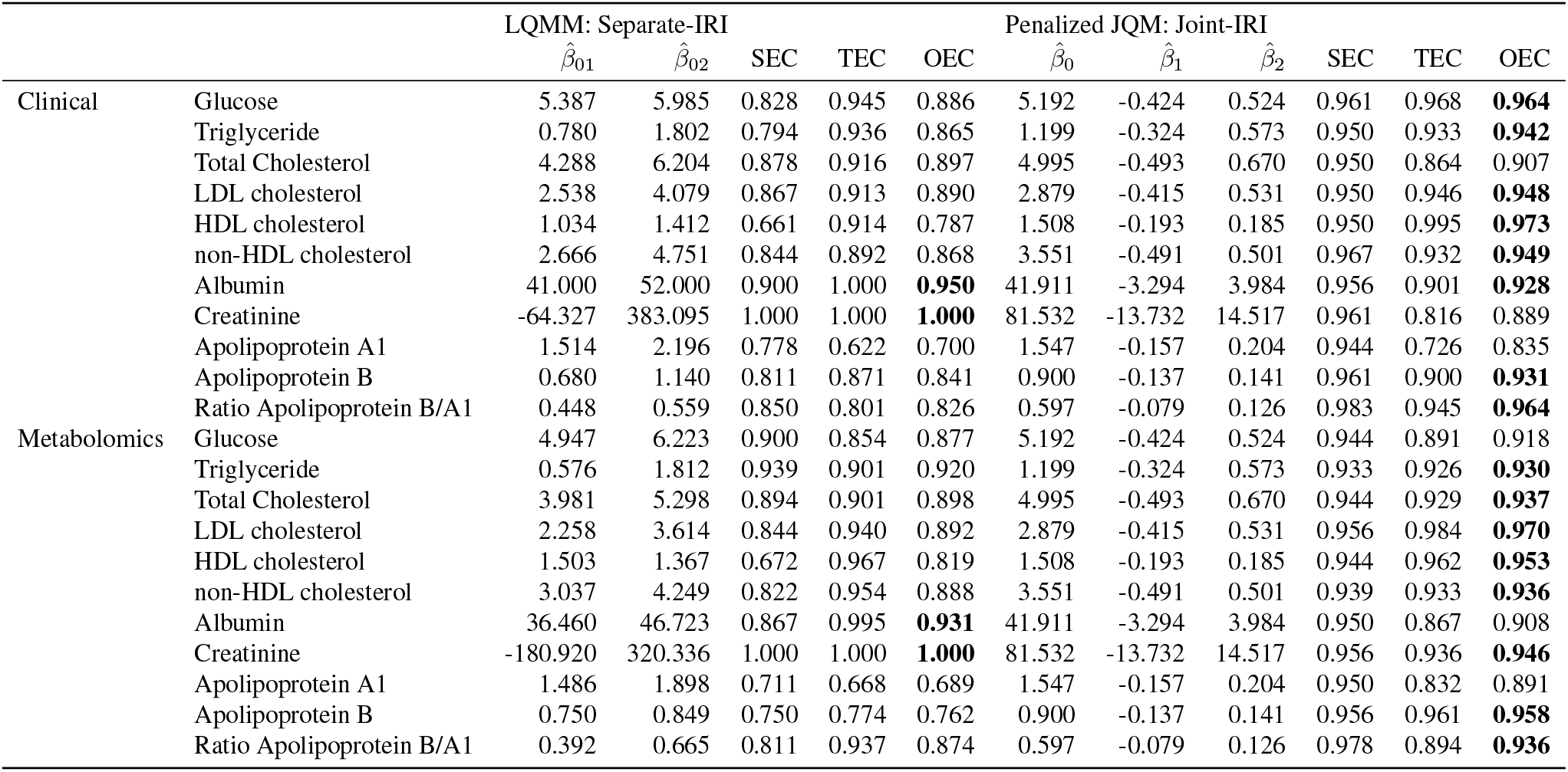
Parameter estimates for the VITO IAM Frontier data. The overall empirical coverages of joint-IRIs are generally higher than for the separate-IRIs, and they are closer to the nominal level (95%). OECs larger than 92.5% are printed in bold face.

We consider the joint-IRIs of glucose and triglyceride from the clinical and metabolomics datasets to discuss some of the features of the IRIs (Figure 5). As before, the IRIs show between subject variability. While for glucose most of the IRIs fall completely within the PRI, some subjects have IRIs that have one or two bounds outside of the PRI (e.g. Subject 29). If it is indeed correct that all subjects in the IAM Frontier dataset are healthy, this is an illustration of a subject for whom large glocuse levels are normal. On the other hand, this subject has several of its measurements outside the PRI and its IRI contains the largest glucose levels (among all subjects in the dataset). This may perhaps be interpreted by the GP as an indication that this subject is at risk for certain diseases. The IRIs for triglyceride show again between-subject variability. Here we can also clearly see the effect of information-sharing, which is a consequence of the estimation method and which is needed to allow for the IRI calculation based on only a few observations per subject. In particular, several subjects have observations that are within the PRI and that show rather small variability (e.g. Subjects 16-20 and Subject 29). However, their IRIs are much wider than suggested by their data; their observations are even not nicely centered within their IRIs. This is a consequence of the information-sharing: some other subjects have much larger triglyceride concentrations (e.g. Subjects 4 and 5) with much larger variability, and this thus also effects the IRIs of the other subjects. This not a problematic feature, because over all subjects the coverage is controlled. Moreover, the IRIs can also be seen as a compromise between the PRI (population interpretation) and the a genuine individual RI that would be estimated from only the subject’s data. Finally, when the number of subjects and the number of repeated measurements within subjects increase, the effects of a few outlying subjects (as e.g. Subject 4) will diminish.

**Figure 5.**
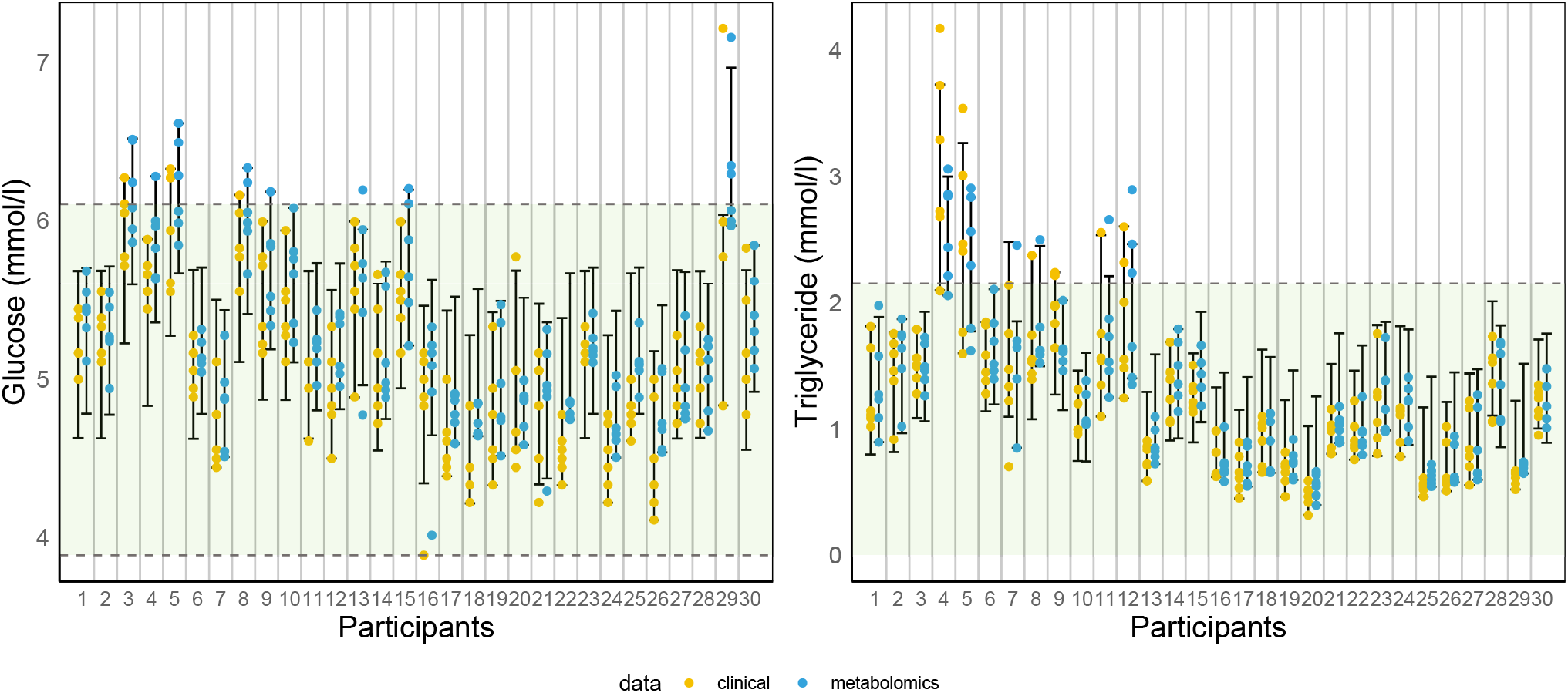
Joint-IRI for all subjects in the VITO IAM Frontier dataset, estimated using the Penalized JQM. Yellow and blue dotted points refer to clinical and metabolomics observations. The green transparent area indicates the published PRI.^12^ The order of individuals is randomised and different between glucose and triglycerides measurements.

## Discussion and Conclusion

We have introduced two individual reference intervals estimation methods that are applicable for the personalized interpretation of clinical and metabolomics outcomes. Linear Quantile Mixed Models (LQMM) serve as a foundation for the first method for constructing the separate-IRI. The variability between the separate-IRIs motivates the need for subject-specific interpretation of reference intervals.^2^ In this paper, we have proposed a Joint Quantile Model (JQM) and a penalized estimation procedure for its parameters. This procedure does not require strong distributional assumptions, in contrast to the LQMM method. Due to the joint modelling of the lower and upper quantiles, and the introduction of penalty parameters, the subject-specific parameters (that specify the position and the length of the IRI) are estimated using all available data. This can also be interpreted as an example of information sharing: even though each subject may only contribute a small number of outcome measurements that is insufficient for the nonparametric estimation of a reference interval, information is shared among subjects, allowing for the estimation of the intervals. Our simulation study demonstrates that for various sample sizes and numbers of repeated outcomes, the joint-IRIs always give empirical coverage values close to the true nominal coverage level, outperforming the separate-IRIs. The performance of the PJQM method is not strongly affected by the shape of outcome distributions. These findings suggest the robustness of the method.

We have illustrated both the separate-IRI and joint-IRI implementation on the clinical and metabolomics data from VITO IAM Frontier. The superiority of the joint-IRI has also been validated in this real-life data application. The overall empirical coverages (OEC) were generally higher in joint-IRIs as compared to the separate-IRIs.

In current practice, clinical decisions are usually binary; if the observation is within the PRI, it is considered normal. If the observation is outside the PRI, an action is usually taken. The IRI range extends this interpretation by providing a personal context to the PRI. For example, if an IRI range is on the PRI boundaries, the individual is likely developing a disease and preventive action should be taken. Whereas if the IRI is clearly within or outside the PRI, the interpretation would be the same. An extended study with different real datasets, i.e. in a larger cohort in healthy and diseased populations, would give a valuable insight into its clinical utility. Furthermore, currently it is a challenge to collect multiple data points from a single individual (which is necessary for calculating IRIs) where his/her data is scattered across various hospitals, institutions and e-health records due to technical difficulties and privacy concerns.^13^ Cloud-based Linked Data solutions are being developed at the European level to address these challenges,^14^ hence enabling data coverage at the individual level.

We believe that when the IRI is widely used in daily clinical practice for interpretation of the results complementary to PRIs, it will be immensely beneficial to extend the use of clinical, metabolomics, and proteomics parameters to precision (and preventive) health.

## Data Availability

Supplementary material of this article is available online. The data that supports the simulation findings are reproducible. The codes to generate the data and produce the results are available online at https://github.com/murihpusparum/PenalizedJQM and in the Supplementary Document. 
The VITO IAM Frontier data is subject to data protection and privacy of data subjects. It is available upon request.

## Acknowledgements

We thank Alejandro Correa Rojo for his technical support in creating the circos plot.

## Data availability

The data that supports the simulation findings are reproducible. The codes to generate the data and produce the results are available online at https://github.com/murihpusparum/PenalizedJQM and in the Supplementary Document. The VITO IAM Frontier data is subject to data protection and privacy of data subjects. It is available upon request.

## Supplemental material

Supplementary material of this article is available online.

