## Supplementary Document for "Individual Reference Intervals for Personalized Interpretation of Clinical and Metabolomics Measurements"

### 1 Parameter Estimation in Penalized JQM

Since the objective function in Section 2.2.2. is a non-convex function, the minimization is normally done by using linear programming methods as in quantile regression. In this study, however, we propose to apply the iterative procedure.

First we describe the estimation procedure with fixed penalty parameters $\lambda_u$ and $\lambda_v$. The estimates of $\beta_0$ and the $u_i$s are computed but before entering the iterative procedure.

#### 1.1 Estimation of $\beta_0$

We estimate $\beta_0$ as the median of all data.

#### 1.2 Initial estimation of $u_i$

We estimate the $u_i$ by only considering the model for the median: $Q_i(0.5) = \beta_0 + u_i$. For this quantile, and with $\beta_0$ already estimated, the objective function reduces to

$$M(\boldsymbol{u}, \lambda_u) = \sum_{i=1}^{N} \sum_{j=1}^{n_i} \rho_{0.5}(y_{ij} - \hat{\beta}_0 - u_i) + \lambda_u \sum_{i=1}^{N} u_i^2.$$

The $u_i$s are now estimated, one by one, by directly minimising this function.

##### 1.2.1 Initial Estimation of $\beta_1$ and $\beta_2$

The parameters $\beta_1$ and $\beta_2$ are initially estimated under the assumption that the lengths of all IRIs are equal, i.e. $z_i = 1$ for all $i = 1, \ldots, N$. Under this restriction, $\beta_1$ and $\beta_2$ are estimated as the sample $\tau_1$ and $\tau_2$ quantiles of $y_{ij} - \hat{\beta}_0 - \hat{u}_i$, respectively.
The next steps are repeated iteratively until convergence.

##### 1.2.2 Estimation of $z_i$

Let

$$\tilde{y}_{ij} = y_{ij} - \hat{\beta}_0 - \hat{u}_i$$

and consider the objective function (note that the penalty term for $u_i$ is dropped because it does not affect the estimation of the $z_i$s)

$$\sum_{k=1}^{2} \sum_{i=1}^{N} \sum_{j=1}^{n_i} \rho_{\tau_k}(\tilde{y}_{ij} - z_i \hat{\beta}_k) + \lambda_z \sum_{i=1}^{N} (z_i - 1)^2.$$

The $z_i$s are estimated, one by one, by direct minimisation of this objective function. This sequential procedure does not necessarily minimise the objective function simultaneously for all $z_i$s, particularly because of the penalty term. As a consequence, the resulting estimates of the $z_i$s are often not well centered about 1 (which is expected because of the form of the penalty term). To remediate this problem, we add a rescaling step. In partciular, we set the rescaled estimates to $\gamma \hat{z}_i$. Note that this rescaling does not affect the bounds of the IRIs if the $\hat{\beta}_1$ and $\hat{\beta}_2$ parameter estimates are rescaled to $\hat{\beta}_1/\gamma$ and $\hat{\beta}_2/\gamma$, respectively. The scaling factor $\gamma$ is obtained by minimising

$$\sum_{k=1}^{2} \sum_{i=1}^{N} \sum_{j=1}^{n_i} \rho_{\tau_k}(\tilde{y}_{ij} - (\gamma \hat{z}_i)(\hat{\beta}_k/\gamma)) + \lambda_z \sum_{i=1}^{N} (\gamma \hat{z}_i - 1)^2.$$

Note that the first term does not depend on $\gamma$. Hence, only the last terms must by minimised. The solution is given by

$$\gamma = \frac{\sum_{i=1}^{N} z_i}{\sum_{i=1}^{N} z_i^2}.$$

The rescaled $z_i$ estimates are again denoted by $\hat{z}_i$.

##### 1.2.3 Estimation of $\beta_1$ and $\beta_2$

First note that there is no penalisation acting on the $\beta$-parameters. With $\tilde{y}_{ij} = y_{ij} - \hat{\beta}_0 - \hat{u}_i$ we write the objective function as

$$D(\beta_1, \beta_2) = \sum_{i=1}^{N} \sum_{j=1}^{n_i} \rho_{\tau_1}(\tilde{y}_{ij} - \hat{z}_i \beta_1) + \sum_{i=1}^{N} \sum_{j=1}^{n_i} \rho_{\tau_2}(\tilde{y}_{ij} - \hat{z}_i \beta_2).$$

Each of the two terms is exactly the objective function of a quantile regression model with $\tilde{y}_{ij}$ as the outcomes and $\hat{z}_i$ as the regressor. Thus the estimates of $\beta_1$ and $\beta_2$ can be obtained by fitting thus quantile regression model.

### 2 Simulation Study

Table S1: Simulation scenarios. The column "Method" indicates what methods were evaluated. A standard normal distribution $N(0,1)$ was used to generate all the error terms in the LQMM method.

| N | n | $\tau_2 - \tau_1$ | $u_i$ | $\varepsilon_{ij}$ | $a$ | $b$ | Method | N | n | $\tau_2 - \tau_1$ | $u_i$ | $\varepsilon_{ij}$ | $a$ | $b$ | Method |
|---|---|---|---|---|---|---|---|---|---|---|---|---|---|---|---|
| 30 | 5 | 0.95 | $N(0,1)$ | $N(0,\theta_i)$ | 2 | 0.4 | LQMM; PJQM | 50 | 10 | 0.90 | scaled$\chi_4^2$ | $N(0,\theta_i)$ | 2 | 0.4 | LQMM; PJQM |
| 30 | 5 | 0.90 | $N(0,1)$ | $N(0,\theta_i)$ | 2 | 0.4 | LQMM; PJQM | 50 | 10 | 0.85 | scaled$\chi_4^2$ | $N(0,\theta_i)$ | 2 | 0.4 | LQMM; PJQM |
| 30 | 5 | 0.85 | $N(0,1)$ | $N(0,\theta_i)$ | 2 | 0.4 | LQMM; PJQM | 50 | 15 | 0.95 | scaled$\chi_4^2$ | $N(0,\theta_i)$ | 2 | 0.4 | LQMM; PJQM |
| 30 | 10 | 0.95 | $N(0,1)$ | $N(0,\theta_i)$ | 2 | 0.4 | LQMM; PJQM | 50 | 15 | 0.90 | scaled$\chi_4^2$ | $N(0,\theta_i)$ | 2 | 0.4 | LQMM; PJQM |
| 30 | 10 | 0.90 | $N(0,1)$ | $N(0,\theta_i)$ | 2 | 0.4 | LQMM; PJQM | 50 | 15 | 0.85 | scaled$\chi_4^2$ | $N(0,\theta_i)$ | 2 | 0.4 | LQMM; PJQM |
| 30 | 10 | 0.85 | $N(0,1)$ | $N(0,\theta_i)$ | 2 | 0.4 | LQMM; PJQM | 50 | 20 | 0.95 | scaled$\chi_4^2$ | $N(0,\theta_i)$ | 2 | 0.4 | LQMM; PJQM |
| 30 | 15 | 0.95 | $N(0,1)$ | $N(0,\theta_i)$ | 2 | 0.4 | LQMM; PJQM | 50 | 20 | 0.90 | scaled$\chi_4^2$ | $N(0,\theta_i)$ | 2 | 0.4 | LQMM; PJQM |
| 30 | 15 | 0.90 | $N(0,1)$ | $N(0,\theta_i)$ | 2 | 0.4 | LQMM; PJQM | 50 | 20 | 0.85 | scaled$\chi_4^2$ | $N(0,\theta_i)$ | 2 | 0.4 | LQMM; PJQM |
| 30 | 15 | 0.85 | $N(0,1)$ | $N(0,\theta_i)$ | 2 | 0.4 | LQMM; PJQM | 30 | 5 | 0.95 | $N(0,1)$ | scaled $t_3$ | 2 | 0.4 | PJQM |
| 30 | 20 | 0.95 | $N(0,1)$ | $N(0,\theta_i)$ | 2 | 0.4 | LQMM; PJQM | 30 | 5 | 0.90 | $N(0,1)$ | scaled $t_3$ | 2 | 0.4 | PJQM |
| 30 | 20 | 0.90 | $N(0,1)$ | $N(0,\theta_i)$ | 2 | 0.4 | LQMM; PJQM | 30 | 5 | 0.85 | $N(0,1)$ | scaled $t_3$ | 2 | 0.4 | PJQM |
| 30 | 20 | 0.85 | $N(0,1)$ | $N(0,\theta_i)$ | 2 | 0.4 | LQMM; PJQM | 30 | 10 | 0.95 | $N(0,1)$ | scaled $t_3$ | 2 | 0.4 | PJQM |
| 50 | 5 | 0.95 | $N(0,1)$ | $N(0,\theta_i)$ | 2 | 0.4 | LQMM; PJQM | 30 | 10 | 0.90 | $N(0,1)$ | scaled $t_3$ | 2 | 0.4 | PJQM |
| 50 | 5 | 0.90 | $N(0,1)$ | $N(0,\theta_i)$ | 2 | 0.4 | LQMM; PJQM | 30 | 10 | 0.85 | $N(0,1)$ | scaled $t_3$ | 2 | 0.4 | PJQM |
| 50 | 5 | 0.85 | $N(0,1)$ | $N(0,\theta_i)$ | 2 | 0.4 | LQMM; PJQM | 30 | 15 | 0.95 | $N(0,1)$ | scaled $t_3$ | 2 | 0.4 | PJQM |
| 50 | 10 | 0.95 | $N(0,1)$ | $N(0,\theta_i)$ | 2 | 0.4 | LQMM; PJQM | 30 | 15 | 0.90 | $N(0,1)$ | scaled $t_3$ | 2 | 0.4 | PJQM |
| 50 | 10 | 0.90 | $N(0,1)$ | $N(0,\theta_i)$ | 2 | 0.4 | LQMM; PJQM | 30 | 15 | 0.85 | $N(0,1)$ | scaled $t_3$ | 2 | 0.4 | PJQM |
| 50 | 10 | 0.85 | $N(0,1)$ | $N(0,\theta_i)$ | 2 | 0.4 | LQMM; PJQM | 30 | 20 | 0.95 | $N(0,1)$ | scaled $t_3$ | 2 | 0.4 | PJQM |
| 50 | 15 | 0.95 | $N(0,1)$ | $N(0,\theta_i)$ | 2 | 0.4 | LQMM; PJQM | 30 | 20 | 0.90 | $N(0,1)$ | scaled $t_3$ | 2 | 0.4 | PJQM |
| 50 | 15 | 0.90 | $N(0,1)$ | $N(0,\theta_i)$ | 2 | 0.4 | LQMM; PJQM | 30 | 20 | 0.85 | $N(0,1)$ | scaled $t_3$ | 2 | 0.4 | PJQM |
| 50 | 15 | 0.85 | $N(0,1)$ | $N(0,\theta_i)$ | 2 | 0.4 | LQMM; PJQM | 50 | 5 | 0.95 | $N(0,1)$ | scaled $t_3$ | 2 | 0.4 | PJQM |
| 50 | 20 | 0.95 | $N(0,1)$ | $N(0,\theta_i)$ | 2 | 0.4 | LQMM; PJQM | 50 | 5 | 0.90 | $N(0,1)$ | scaled $t_3$ | 2 | 0.4 | PJQM |
| 50 | 20 | 0.90 | $N(0,1)$ | $N(0,\theta_i)$ | 2 | 0.4 | LQMM; PJQM | 50 | 5 | 0.85 | $N(0,1)$ | scaled $t_3$ | 2 | 0.4 | PJQM |
| 50 | 20 | 0.85 | $N(0,1)$ | $N(0,\theta_i)$ | 2 | 0.4 | LQMM; PJQM | 50 | 10 | 0.95 | $N(0,1)$ | scaled $t_3$ | 2 | 0.4 | PJQM |
| 30 | 5 | 0.95 | scaled$\chi_4^2$ | $N(0,\theta_i)$ | 2 | 0.4 | LQMM; PJQM | 50 | 10 | 0.90 | $N(0,1)$ | scaled $t_3$ | 2 | 0.4 | PJQM |
| 30 | 5 | 0.90 | scaled$\chi_4^2$ | $N(0,\theta_i)$ | 2 | 0.4 | LQMM; PJQM | 50 | 10 | 0.85 | $N(0,1)$ | scaled $t_3$ | 2 | 0.4 | PJQM |
| 30 | 5 | 0.85 | scaled$\chi_4^2$ | $N(0,\theta_i)$ | 2 | 0.4 | LQMM; PJQM | 50 | 15 | 0.95 | $N(0,1)$ | scaled $t_3$ | 2 | 0.4 | PJQM |
| 30 | 10 | 0.95 | scaled$\chi_4^2$ | $N(0,\theta_i)$ | 2 | 0.4 | LQMM; PJQM | 50 | 15 | 0.90 | $N(0,1)$ | scaled $t_3$ | 2 | 0.4 | PJQM |
| 30 | 10 | 0.90 | scaled$\chi_4^2$ | $N(0,\theta_i)$ | 2 | 0.4 | LQMM; PJQM | 50 | 15 | 0.85 | $N(0,1)$ | scaled $t_3$ | 2 | 0.4 | PJQM |
| 30 | 10 | 0.85 | scaled$\chi_4^2$ | $N(0,\theta_i)$ | 2 | 0.4 | LQMM; PJQM | 50 | 20 | 0.95 | $N(0,1)$ | scaled $t_3$ | 2 | 0.4 | PJQM |
| 30 | 15 | 0.95 | scaled$\chi_4^2$ | $N(0,\theta_i)$ | 2 | 0.4 | LQMM; PJQM | 50 | 20 | 0.90 | $N(0,1)$ | scaled $t_3$ | 2 | 0.4 | PJQM |
| 30 | 15 | 0.90 | scaled$\chi_4^2$ | $N(0,\theta_i)$ | 2 | 0.4 | LQMM; PJQM | 50 | 20 | 0.85 | $N(0,1)$ | scaled $t_3$ | 2 | 0.4 | PJQM |
| 30 | 15 | 0.85 | scaled$\chi_4^2$ | $N(0,\theta_i)$ | 2 | 0.4 | LQMM; PJQM | 30 | 10 | 0.95 | $N(0,1)$ | $N(0,\theta_i)$ | 2 | 0.8 | PJQM |
| 30 | 20 | 0.95 | scaled$\chi_4^2$ | $N(0,\theta_i)$ | 2 | 0.4 | LQMM; PJQM | 30 | 10 | 0.95 | $N(0,1)$ | $N(0,\theta_i)$ | 2 | 1.6 | PJQM |
| 30 | 20 | 0.90 | scaled$\chi_4^2$ | $N(0,\theta_i)$ | 2 | 0.4 | LQMM; PJQM | 30 | 10 | 0.95 | $N(0,1)$ | $N(0,\theta_i)$ | 2.4 | 0.4 | PJQM |
| 30 | 20 | 0.85 | scaled$\chi_4^2$ | $N(0,\theta_i)$ | 2 | 0.4 | LQMM; PJQM | 30 | 10 | 0.95 | $N(0,1)$ | $N(0,\theta_i)$ | 2.4 | 0.8 | PJQM |
| 50 | 5 | 0.95 | scaled$\chi_4^2$ | $N(0,\theta_i)$ | 2 | 0.4 | LQMM; PJQM | 30 | 10 | 0.95 | $N(0,1)$ | $N(0,\theta_i)$ | 2.4 | 1.6 | PJQM |
| 50 | 5 | 0.90 | scaled$\chi_4^2$ | $N(0,\theta_i)$ | 2 | 0.4 | LQMM; PJQM | 30 | 10 | 0.95 | $N(0,1)$ | $N(0,\theta_i)$ | 2.8 | 0.4 | PJQM |
| 50 | 5 | 0.85 | scaled$\chi_4^2$ | $N(0,\theta_i)$ | 2 | 0.4 | LQMM; PJQM | 30 | 10 | 0.95 | $N(0,1)$ | $N(0,\theta_i)$ | 2.8 | 0.8 | PJQM |
| 50 | 10 | 0.95 | scaled$\chi_4^2$ | $N(0,\theta_i)$ | 2 | 0.4 | LQMM; PJQM | 30 | 10 | 0.95 | $N(0,1)$ | $N(0,\theta_i)$ | 2.8 | 1.6 | PJQM |

Table S2: Estimates of penalty terms $\lambda_u$ and $\lambda_z$ in the PJQM method over different simulation scenarios.

| N | n | $\tau_2 - \tau_1$ | $u_i$ | $\varepsilon_{ij}$ | $a$ | $b$ | $\hat{\lambda}_u$ | $\hat{\lambda}_z$ | N | n | $\tau_2 - \tau_1$ | $u_i$ | $\varepsilon_{ij}$ | $a$ | $b$ | $\hat{\lambda}_u$ | $\hat{\lambda}_z$ |
|---|---|---|---|---|---|---|---|---|---|---|---|---|---|---|---|---|---|
| 30 | 5 | 0.95 | $N(0,1)$ | $N(0,\theta_i)$ | 2 | 0.4 | 1.5 | 2.6 | 50 | 10 | 0.90 | scaled$\chi_4^2$ | $N(0,\theta_i)$ | 2 | 0.4 | 1.4 | 2.5 |
| 30 | 5 | 0.90 | $N(0,1)$ | $N(0,\theta_i)$ | 2 | 0.4 | 1.5 | 2.5 | 50 | 10 | 0.85 | scaled$\chi_4^2$ | $N(0,\theta_i)$ | 2 | 0.4 | 1.4 | 2.5 |
| 30 | 5 | 0.85 | $N(0,1)$ | $N(0,\theta_i)$ | 2 | 0.4 | 1.4 | 2.5 | 50 | 15 | 0.95 | scaled$\chi_4^2$ | $N(0,\theta_i)$ | 2 | 0.4 | 1.4 | 2.6 |
| 30 | 10 | 0.95 | $N(0,1)$ | $N(0,\theta_i)$ | 2 | 0.4 | 1.5 | 2.4 | 50 | 15 | 0.90 | scaled$\chi_4^2$ | $N(0,\theta_i)$ | 2 | 0.4 | 1.3 | 2.4 |
| 30 | 10 | 0.90 | $N(0,1)$ | $N(0,\theta_i)$ | 2 | 0.4 | 1.4 | 2.3 | 50 | 15 | 0.85 | scaled$\chi_4^2$ | $N(0,\theta_i)$ | 2 | 0.4 | 1.3 | 2.5 |
| 30 | 10 | 0.85 | $N(0,1)$ | $N(0,\theta_i)$ | 2 | 0.4 | 1.4 | 2.3 | 50 | 20 | 0.95 | scaled$\chi_4^2$ | $N(0,\theta_i)$ | 2 | 0.4 | 1.3 | 2.7 |
| 30 | 15 | 0.95 | $N(0,1)$ | $N(0,\theta_i)$ | 2 | 0.4 | 1.4 | 2.4 | 50 | 20 | 0.90 | scaled$\chi_4^2$ | $N(0,\theta_i)$ | 2 | 0.4 | 1.2 | 2.5 |
| 30 | 15 | 0.90 | $N(0,1)$ | $N(0,\theta_i)$ | 2 | 0.4 | 1.2 | 2.4 | 50 | 20 | 0.85 | scaled$\chi_4^2$ | $N(0,\theta_i)$ | 2 | 0.4 | 1.2 | 2.6 |
| 30 | 15 | 0.85 | $N(0,1)$ | $N(0,\theta_i)$ | 2 | 0.4 | 1.2 | 2.4 | 30 | 5 | 0.95 | $N(0,1)$ | scaled $t_3$ | 2 | 0.4 | 1.5 | 2.5 |
| 30 | 20 | 0.95 | $N(0,1)$ | $N(0,\theta_i)$ | 2 | 0.4 | 1.3 | 2.6 | 30 | 5 | 0.90 | $N(0,1)$ | scaled $t_3$ | 2 | 0.4 | 1.6 | 2.4 |
| 30 | 20 | 0.90 | $N(0,1)$ | $N(0,\theta_i)$ | 2 | 0.4 | 1.2 | 2.3 | 30 | 5 | 0.85 | $N(0,1)$ | scaled $t_3$ | 2 | 0.4 | 1.4 | 2.5 |
| 30 | 20 | 0.85 | $N(0,1)$ | $N(0,\theta_i)$ | 2 | 0.4 | 1.2 | 2.5 | 30 | 10 | 0.95 | $N(0,1)$ | scaled $t_3$ | 2 | 0.4 | 1.6 | 2.2 |
| 50 | 5 | 0.95 | $N(0,1)$ | $N(0,\theta_i)$ | 2 | 0.4 | 1.6 | 2.7 | 30 | 10 | 0.90 | $N(0,1)$ | scaled $t_3$ | 2 | 0.4 | 1.5 | 2.1 |
| 50 | 5 | 0.90 | $N(0,1)$ | $N(0,\theta_i)$ | 2 | 0.4 | 1.5 | 2.7 | 30 | 10 | 0.85 | $N(0,1)$ | scaled $t_3$ | 2 | 0.4 | 1.5 | 2.2 |
| 50 | 5 | 0.85 | $N(0,1)$ | $N(0,\theta_i)$ | 2 | 0.4 | 1.4 | 2.5 | 30 | 15 | 0.95 | $N(0,1)$ | scaled $t_3$ | 2 | 0.4 | 1.3 | 2.2 |
| 50 | 10 | 0.95 | $N(0,1)$ | $N(0,\theta_i)$ | 2 | 0.4 | 1.5 | 2.6 | 30 | 15 | 0.90 | $N(0,1)$ | scaled $t_3$ | 2 | 0.4 | 1.3 | 2.1 |
| 50 | 10 | 0.90 | $N(0,1)$ | $N(0,\theta_i)$ | 2 | 0.4 | 1.5 | 2.4 | 30 | 15 | 0.85 | $N(0,1)$ | scaled $t_3$ | 2 | 0.4 | 1.3 | 2.2 |
| 50 | 10 | 0.85 | $N(0,1)$ | $N(0,\theta_i)$ | 2 | 0.4 | 1.4 | 2.3 | 30 | 20 | 0.95 | $N(0,1)$ | scaled $t_3$ | 2 | 0.4 | 1.3 | 1.8 |
| 50 | 15 | 0.95 | $N(0,1)$ | $N(0,\theta_i)$ | 2 | 0.4 | 1.5 | 2.5 | 30 | 20 | 0.90 | $N(0,1)$ | scaled $t_3$ | 2 | 0.4 | 1.0 | 2.0 |
| 50 | 15 | 0.90 | $N(0,1)$ | $N(0,\theta_i)$ | 2 | 0.4 | 1.3 | 2.3 | 30 | 20 | 0.85 | $N(0,1)$ | scaled $t_3$ | 2 | 0.4 | 1.2 | 2.4 |
| 50 | 15 | 0.85 | $N(0,1)$ | $N(0,\theta_i)$ | 2 | 0.4 | 1.2 | 2.4 | 50 | 5 | 0.95 | $N(0,1)$ | scaled $t_3$ | 2 | 0.4 | 1.6 | 2.4 |
| 50 | 20 | 0.95 | $N(0,1)$ | $N(0,\theta_i)$ | 2 | 0.4 | 1.4 | 2.8 | 50 | 5 | 0.90 | $N(0,1)$ | scaled $t_3$ | 2 | 0.4 | 1.5 | 2.5 |
| 50 | 20 | 0.90 | $N(0,1)$ | $N(0,\theta_i)$ | 2 | 0.4 | 1.2 | 2.6 | 50 | 5 | 0.85 | $N(0,1)$ | scaled $t_3$ | 2 | 0.4 | 1.5 | 2.2 |
| 50 | 20 | 0.85 | $N(0,1)$ | $N(0,\theta_i)$ | 2 | 0.4 | 1.1 | 2.5 | 50 | 10 | 0.95 | $N(0,1)$ | scaled $t_3$ | 2 | 0.4 | 1.6 | 2.1 |
| 30 | 5 | 0.95 | scaled$\chi_4^2$ | $N(0,\theta_i)$ | 2 | 0.4 | 1.4 | 2.7 | 50 | 10 | 0.90 | $N(0,1)$ | scaled $t_3$ | 2 | 0.4 | 1.7 | 2.2 |
| 30 | 5 | 0.90 | scaled$\chi_4^2$ | $N(0,\theta_i)$ | 2 | 0.4 | 1.5 | 2.6 | 50 | 10 | 0.85 | $N(0,1)$ | scaled $t_3$ | 2 | 0.4 | 1.6 | 2.1 |
| 30 | 5 | 0.85 | scaled$\chi_4^2$ | $N(0,\theta_i)$ | 2 | 0.4 | 1.4 | 2.5 | 50 | 15 | 0.95 | $N(0,1)$ | scaled $t_3$ | 2 | 0.4 | 1.4 | 1.7 |
| 30 | 10 | 0.95 | scaled$\chi_4^2$ | $N(0,\theta_i)$ | 2 | 0.4 | 1.4 | 2.6 | 50 | 15 | 0.90 | $N(0,1)$ | scaled $t_3$ | 2 | 0.4 | 1.4 | 1.9 |
| 30 | 10 | 0.90 | scaled$\chi_4^2$ | $N(0,\theta_i)$ | 2 | 0.4 | 1.4 | 2.4 | 50 | 15 | 0.85 | $N(0,1)$ | scaled $t_3$ | 2 | 0.4 | 1.3 | 2.1 |
| 30 | 10 | 0.85 | scaled$\chi_4^2$ | $N(0,\theta_i)$ | 2 | 0.4 | 1.3 | 2.4 | 50 | 20 | 0.95 | $N(0,1)$ | scaled $t_3$ | 2 | 0.4 | 1.1 | 1.5 |
| 30 | 15 | 0.95 | scaled$\chi_4^2$ | $N(0,\theta_i)$ | 2 | 0.4 | 1.4 | 2.5 | 50 | 20 | 0.90 | $N(0,1)$ | scaled $t_3$ | 2 | 0.4 | 1.0 | 1.8 |
| 30 | 15 | 0.90 | scaled$\chi_4^2$ | $N(0,\theta_i)$ | 2 | 0.4 | 1.2 | 2.5 | 50 | 20 | 0.85 | $N(0,1)$ | scaled $t_3$ | 2 | 0.4 | 1.1 | 2.1 |
| 30 | 15 | 0.85 | scaled$\chi_4^2$ | $N(0,\theta_i)$ | 2 | 0.4 | 1.2 | 2.4 | 30 | 10 | 0.95 | $N(0,1)$ | $N(0,\theta_i)$ | 2.4 | 0.4 | 1.5 | 2.3 |
| 30 | 20 | 0.95 | scaled$\chi_4^2$ | $N(0,\theta_i)$ | 2 | 0.4 | 1.3 | 2.6 | 30 | 10 | 0.95 | $N(0,1)$ | $N(0,\theta_i)$ | 2.8 | 0.4 | 1.5 | 2.3 |
| 30 | 20 | 0.90 | scaled$\chi_4^2$ | $N(0,\theta_i)$ | 2 | 0.4 | 1.2 | 2.5 | 30 | 10 | 0.95 | $N(0,1)$ | $N(0,\theta_i)$ | 2 | 0.8 | 1.4 | 2.6 |
| 30 | 20 | 0.85 | scaled$\chi_4^2$ | $N(0,\theta_i)$ | 2 | 0.4 | 1.2 | 2.5 | 30 | 10 | 0.95 | $N(0,1)$ | $N(0,\theta_i)$ | 2.4 | 0.8 | 1.5 | 2.5 |
| 50 | 5 | 0.95 | scaled$\chi_4^2$ | $N(0,\theta_i)$ | 2 | 0.4 | 1.5 | 2.8 | 30 | 10 | 0.95 | $N(0,1)$ | $N(0,\theta_i)$ | 2.8 | 0.8 | 1.4 | 2.4 |
| 50 | 5 | 0.90 | scaled$\chi_4^2$ | $N(0,\theta_i)$ | 2 | 0.4 | 1.5 | 2.7 | 30 | 10 | 0.95 | $N(0,1)$ | $N(0,\theta_i)$ | 2 | 1.6 | 1.2 | 2.9 |
| 50 | 5 | 0.85 | scaled$\chi_4^2$ | $N(0,\theta_i)$ | 2 | 0.4 | 1.4 | 2.5 | 30 | 10 | 0.95 | $N(0,1)$ | $N(0,\theta_i)$ | 2.4 | 1.6 | 1.3 | 2.8 |
| 50 | 10 | 0.95 | scaled$\chi_4^2$ | $N(0,\theta_i)$ | 2 | 0.4 | 1.5 | 2.7 | 30 | 10 | 0.95 | $N(0,1)$ | $N(0,\theta_i)$ | 2.8 | 1.6 | 1.4 | 2.6 |

### 3 IAM Frontier Data

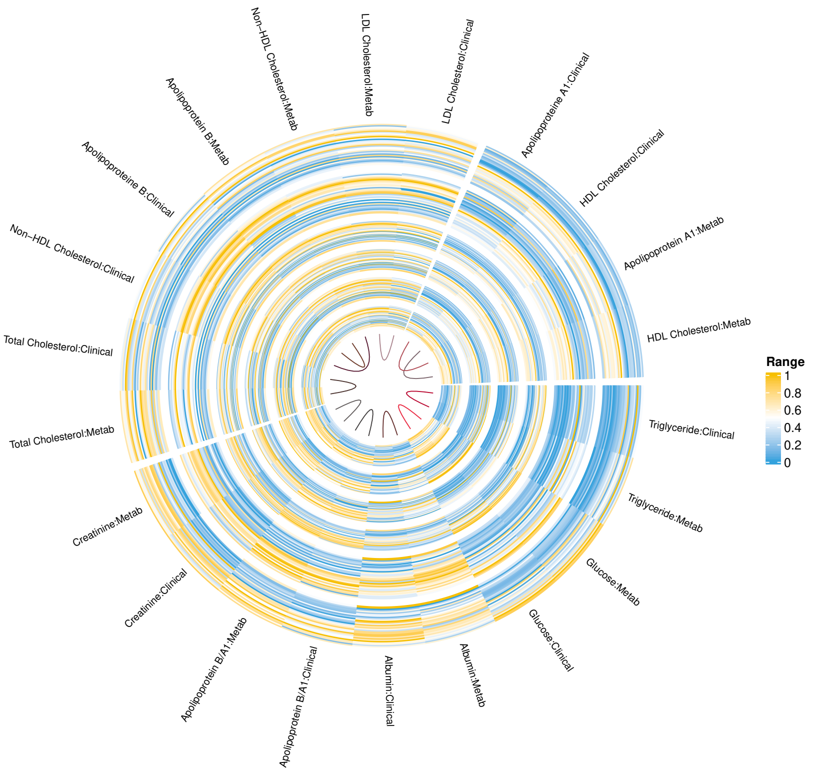


Figure S1: Circos plot showing an overview of the VITO IAM Frontier clinical and metabolomics data over six time-points for thirty participants. All data is converted to the same units and Min-Max scaled for visualization where orange represent the highest and blue represent the lowest value.

We present a general overview of the data in Figure S1. The figure shows different variables as independent arcs on each circle (annulus) where each circle consists of thirty individuals data for this time point. Six circles represent the time points that are collected and the bands show the similarity of each variable to another within the complete dataset. The creation of the bands were unsupervised and their perfect matching of each clinical parameter to its metabolomics counterpart points out to the reproducibility of these measurements by both clinical and metabolomics technologies. Data is demonstrated by using three clusters ($k$-means clustering; $k = 3$) where the variables within each cluster corresponds to both biologically codependent processes (such as Apolipoprotein A1 is the major protein component of HDL particles in plasma hence the two variables are dependent).

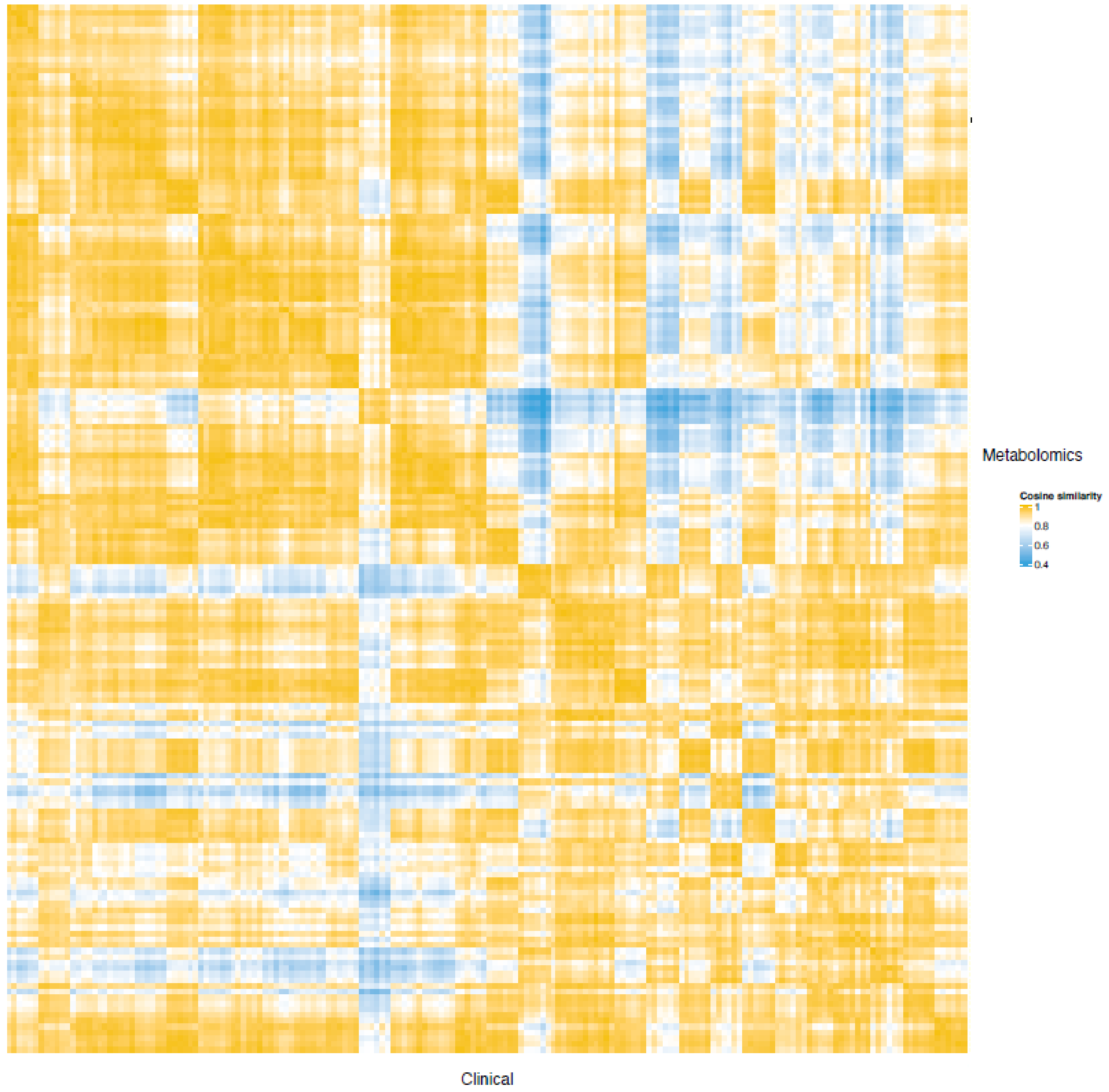


Figure S2: Cosine similarity matrix between eleven shared clinical ($x$-axis) and metabolomics ($y$-axis) parameters computed from IAM Frontier data. The darker yellow color in the diagonal shows the higher similarity between clinical and metabolomics outcome. Overall, the similarity are very high between measurements from the same sample; with a median of 0.982, minimum of 0.881 and maximum of 0.997.

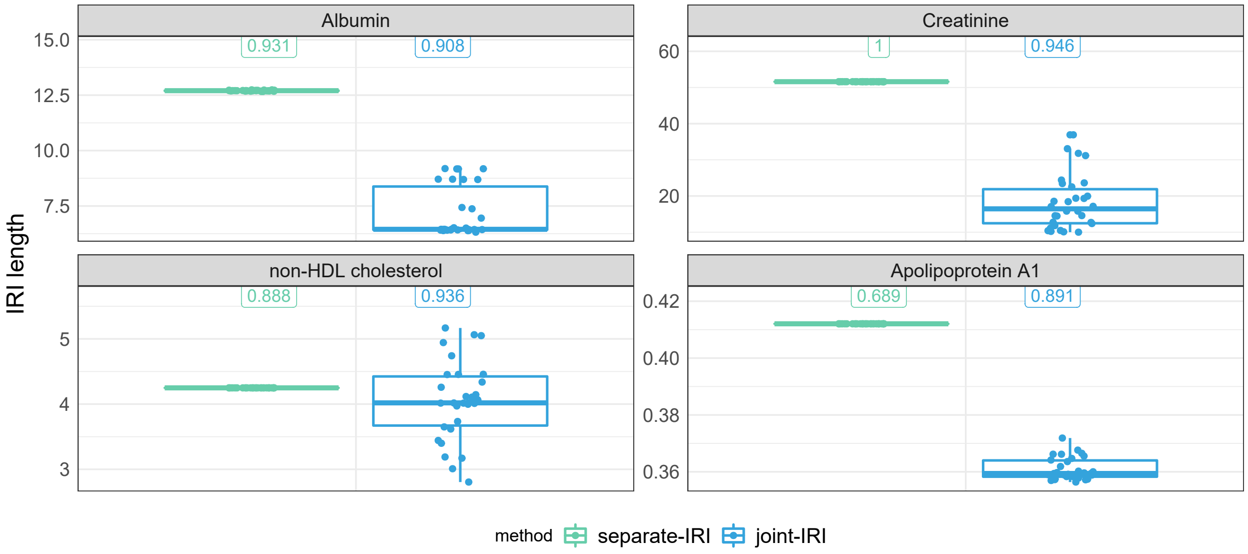


Figure S3: The IRI length distributions of four outcomes measured in the VITO IAM Frontier metabolomics data. The green and blue boxplots refer to IRI lengths estimated by the LQMM method (separate-IRI) and the PJQM method (joint-IRI), respectively. The values on top of each boxplot refer to the corresponding OECs. Extremely small variabilities were observed on the length of separate-IRI due to the limitation of the algorithm to converge at the extreme value of quantile level (e.g. 0.975 or 0.025).

### 4 IAM Frontier Data Description

Metabolite quantification Metabolites were quantified from EDTA plasma samples of 30 individuals, analyzed using the same high-throughput H-NMR metabolomics platform (Nightingale Health Ltd., Helsinki, Finland; nightingalehealth.com/).

Clinical Laboratory Tests: Clinical laboratory tests from blood plasma samples are performed by a certified Clinical laboratory (Algemeen Medisch Laboratorium – AML in Antwerpen, Belgium; www.aml-lab.be).
